# Prevalence and mortality of Lung Comorbidities Among Patients with COVID-19: A systematic review and meta-analysis

**DOI:** 10.1101/2020.06.01.20119271

**Authors:** Mohammed G Alkhathami, Shailesh M Advani, Adil A Abalkhail, Fahad M Alkhathami, Mohammed K Alshehri, Ebtisam Albeashy, Jihad A Alsalamah

## Abstract

**Background:** COVID-19 infections are seen across all age groups but they have shown to have a predisposition for the elderly and those with underlying comorbidities. Patients with severe COVID-19 infections and comorbidities are more prone to respiratory distress syndrome (ARDS), mechanical ventilator use and ultimately succumb to these complications. Little evidence exists of the prevalence of underlying lung comorbidities among COVID-19 patients and associated mortality.

**Methods:** We performed a systematic review of the literature including PubMed (Medline), Embase (Ovid), Google Scholar and Cochrane Library. The last date for our search was 29th April 2020. We included all original research articles on COVID-19 and calculated prevalence of chronic lung disease patients among COVID-19 patients using random effects model. Further we assessed for mortality rates among COVID-19 patients associated with these lung comorbidities.

**Results:** The authors identified 29 articles that reported prevalence of chronic lung conditions among COVID-19 patients. Among those, 26 were from China and 3 from the United States. The pooled prevalence of lung comorbidities including Asthma, COPD, and lung cancer was 3% (95% CI=0-14%), 2.2% (95% CI=0.02-0.03%) and 2.1% (95% CI=0.00-0.21%) respectively. Mortality rates associated with these comorbidities was 30% (41/137) for COPD and 19% (7/37) for lung cancer respectively. No mortality rates were reported for patients with asthma.

**Conclusion:** This study offers latest evidence of prevalence of chronic lung conditions among patients with COVID-19. Asthma, followed by COPD and lung cancer, was the most common lung comorbidity associated with COVID-19, while the higher mortality rate was found in COPD. Future studies are needed to assess other lung comorbidities and associated mortality among patients diagnosed with COVID-19.

## Introduction

The novel coronavirus (2019-nCoV/SARS-Cov-2) also termed as COVID-19 discovered in late 2019 in Wuhan, China has been established to be the main cause of a cluster of pneumonia cases which have disrupted the world order today.^1^. As of May 26th, 2019, the spread of COVID-19 has been rapid across the globe with more than 5,307,298 confirmed cases of COVID-19, including 342,070 deaths, have been reported by the World Health Organization. Given the rapid spread of COVID-19 and driven by international concern, the WHO declared coronavirus as a global pandemic.^2^ Caused by a new strain of the virus, COVID-19 belongs to the family of the class of coronavirus that included viruses associated with the outbreak of Middle East Respiratory (MERS) in 2012 and severe acute respiratory syndrome (SARS) in 2002.^3^. Today, most COVID-19 patients show symptoms of viral pneumonia such as fever, fatigue, dry cough, and lymphopenia.^4^

Reports from around the world highlight that COVID-19 infection is more common among the elderly, those who live in elderly homes or nursing homes, and those with preexisting conditions like diabetes, heart diseases, and cancer.^5^ Rapidly evolving evidence highlight the burden and rates of infection among people with and without comorbidities and see a significant higher burden among those with comorbidities.^6^ Especially, in this early systematic review and meta-analysis, authors identified that most prevalent comorbidities among patients with COVID-19 included hypertension (21.1%, 95% CI: 13.0–27.2%), diabetes (9.7%, 95% CI: 7.2–12.2%), cardiovascular disease (8.4%, 95% CI: 3.8–13.8%) and respiratory system disease (1.5%, 95% CI: 0.9–2.1%).^6^ In addition to symptoms associated with pneumonia, COVID-19 has been shown to cause damage to other organs including heart, liver, and kidneys as well as blood and immune system.^4^ In severe COVID-19 infection, patients develop or succumb to multiple organ failure, shock, acute respiratory distress syndrome, heart failure, arrhythmias, and renal failure.^7^

Finally, smoking and associated comorbidities have shown to increase the risk of infection with COVID-19 due to underlying mechanisms surrounding angiotensin-converting enzyme receptors and high affinity of COVID-19 to ACE receptors.^8^ According to recent findings, COVID-19 related lung injury leading to acute respiratory distress syndrome remains the leading cause of mortality worldwide.^9^ This includes the development of secondary haemophagocytic lymphohistiocytosis (sHLH), a hyperinflammatory syndrome triggered by a viral infection and characterized by a fulminant and fatal hypercytokinaemia with multiorgan failure and ARDS is the hallmark of COVID-19 infection-associated mortality.^9^ Due to the underlying predisposition of COVID-19 to lung tissue, it remains imperative to assess the burden of lung-specific comorbidities including COPD, asthma, lung cancer, and cystic fibrosis among COVID-19 patients. This remains crucial to improving outcomes among those with underlying comorbidities as they might share common underlying mechanisms. Up to date, there is no comprehensive evidence to evaluate the prevalence of chronic lung diseases in COVID-19 patients. Therefore, the primary objective of this systemic review and meta-analysis is to determine the prevalence and mortality of chronic lung diseases which include COPD, lung cancer, asthma, and cystic fibrosis in confirmed COVID-19 cases.

## Methods

### Search Strategy

We registered this systematic review with PROSPERO (ID: CRD42020178039). Also, it was conducted in accordance with the Preferred Reporting Items for Systematic Reviews and Meta-Analyses (PRISMA) guidelines.^10^

Authors performed a literature search of scientific databases including PubMed (Medline), Embase (Ovid), and the Cochrane Library. Furthermore, we did an additional search on Google Scholar. The date of our search was from inception to April 29th, 2020. Keywords used for our search included: “COVID-19 AND chronic lung disease” OR “COVID-19 AND Comorbidities “ OR “new coronavirus AND Characteristics” OR “ new coronavirus AND Comorbidities” OR “Coronavirus AND characteristics AND Comorbidities” OR “Coronavirus Infections” OR “ SARS-CoV-2 AND characteristics” OR “ SARS-CoV-2 AND Comorbidities”). The search strategy was developed for one database and then modified for other search engines. Furthermore, we did an additional search from references of included studies and published preprints. Standard excel sheets were used for data import, abstract screening and full text review. Two reviewers independently screened all abstracts. They rated abstracts as yes or no for inclusion in full text review. For those abstracts, where a decision could not be reached, decision was reached through mutual consensus.

### Inclusion and Exclusion Criteria

We included all studies in English language that were peer-reviewed and preprints of patients diagnosed with COVID-19 and incorporated information on lung comorbidities including Asthma, COPD, Lung cancer, and Cystic fibrosis. We excluded those that did not report on these conditions, not full text, were reviews articles or meta-analysis, perspectives, comments, and other reports.

### Data Extraction and Paper Quality Evaluation

Two authors (M.A. and J.A.) screened and evaluated the literature independently for both narrative synthesis and statistical analysis. From each study, various details including the authors’ names, study design/country, study population, age, and acknowledged features and mortality of Asthma, COPD, Lung cancer and Cystic fibrosis, extracted into Microsoft excel sheet. Detailed description and characteristics of included studies are summarized in Table 1. Moreover, two independent authors were assessed all the included papers using the modified version of the Newcastle-Ottawa Scale (NOS) and the results are provided in Table 1.^11^ This detailed table includes seven domains, each one of these domains was scored from 0 (high risk of bias) to 1 (low risk of bias) and we took a total of the domains to result in a score between 0 to 7, where a higher score indicates a low risk of bias.

**Table 1:** Characteristics of Included Studies on Lung Comorbidities among COVID-19 patients

### Statistical Analysis

During this collective investigation, we calculated pooled estimates of chronic lung conditions among COVID-19 patients. We used random effects model to estimate these prevalence’s among COVID-19 patients. Further we assessed for heterogeneity and publication bias. In addition, 95% confidence interval used to estimate the Inverse variance method, and heterogeneity was evaluated by using I^^^2. Further, the restricted maximum-likelihood estimator was used to estimate the between-study variance biases. Data were displayed using forest plots and all statistical analyses were performed using metaprop command in R metaprop command. Further publication bias was assessed using Funnel Plots and Egger’s regression test.^12, 13^

#### Steps involved in Publication bias

We used the trim-and-fill procedure to account for publication bias.^14^ It includes the following five steps:

a. Estimating the number of studies in the outlying (right) part of the funnel plot.
b. Removing (trimming) these effect sizes and pooling the results with the remaining effect sizes.
c. This pooled effect is then taken as the center of all effect sizes.
d. For each trimmed/removed study, an additional study is imputed, mirroring the effect of the study on the left side of the funnel plot.
e. Pooling the results with the imputed studies and the trimmed studies included.

## Result

### Characteristics of included studies

A summary of the included studies is presented in Table 1, which included a total of 6261 confirmed COVID-19 patients across 29 included studies. Majority of our participants were males (54%). Due to the novel nature and outbreak of COVID-19, our articles found that only three lung comorbidities were included in these articles. These included asthma, COPD, and lung cancer respectively. Surprisingly, no studies estimated prevalence of cystic fibrosis that increases risk of individuals to opportunistic infections. This might be due to it being a rare disease and having low population prevalence. The sample size of the included studies ranged from 2 to 1590 patients. Most of the studies were retrospective in nature and based mainly in China (n=26) followed by the United States (n=3). Further mortality rates were only reported for COPD and lung cancer. The crude mortality rate was 30% (41/137) and 19% (7/37), respectively. Figure 1 provides an overview of our selection process using the PRISMA flow chart.

**Fig 1.**
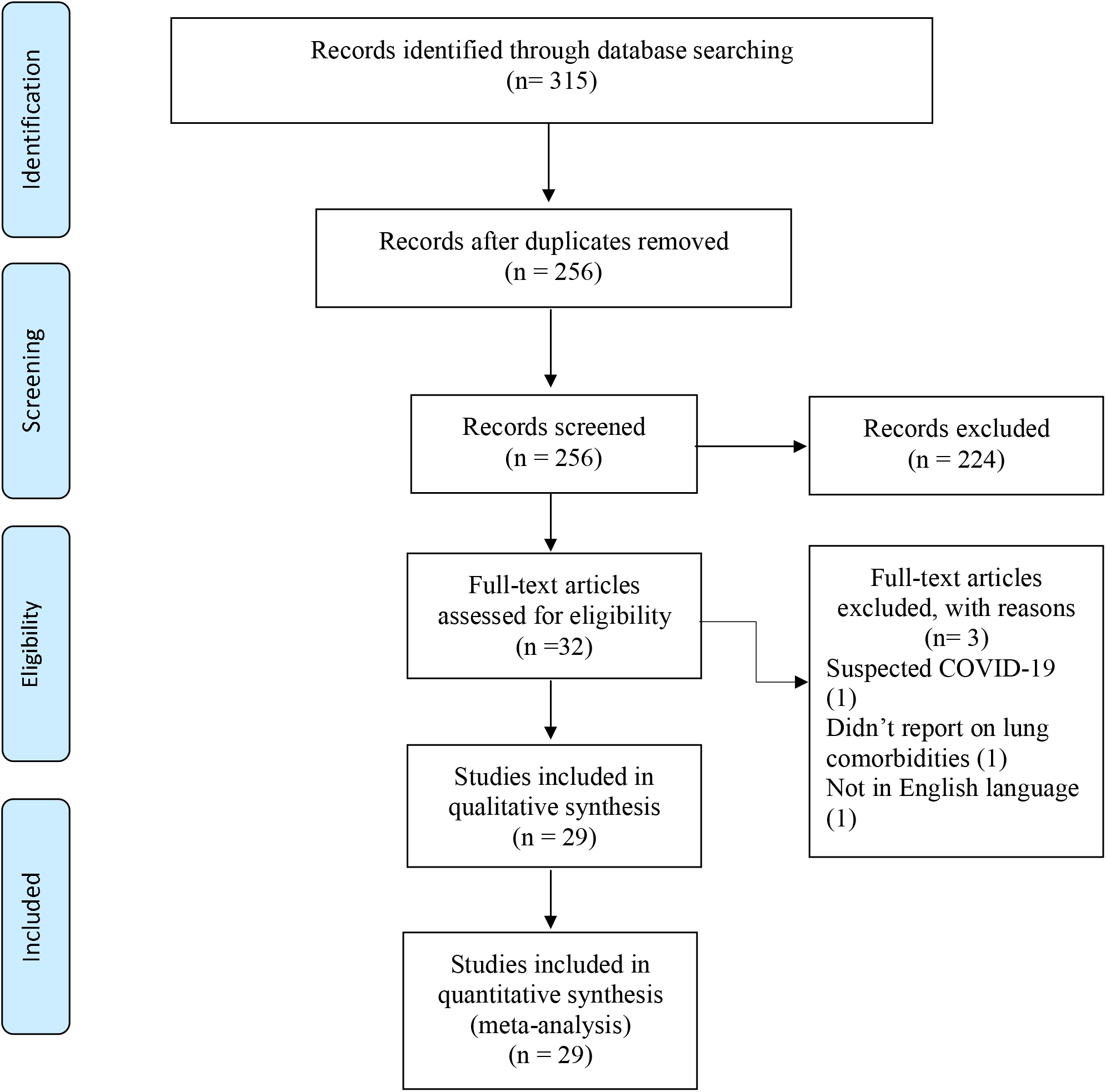
PRISMA flow chart of the systematic literature review and article identification.

### Meta-analysis of prevalence of lung comorbidities among patients with COVID-19

## Asthma

We identified 8 studies that measured prevalence of asthma among patients diagnosed with COVID-19.^15-22^ In pooled analysis we found that prevalence of asthma among patients hospitalized with COVID-19 was 3% (range 0-14%). Figure 2 shows the forest plot for asthma prevalence among COVID-19 patients. Our analysis identified significant presence of heterogeneity in our estimate (*I*^2^ =79%, p<0.01). There was significant publication bias in studies was present because there was asymmetry of the funnel plot presented in figure 3 based on Egger’s test was (t=2.71, p=0.04). Therefore, we performed a trim and fill method in figure 3. This estimate showed little change, which indicated influence of the publication bias was small (t = 0.15, p = 0.89).

**Fig 2:**
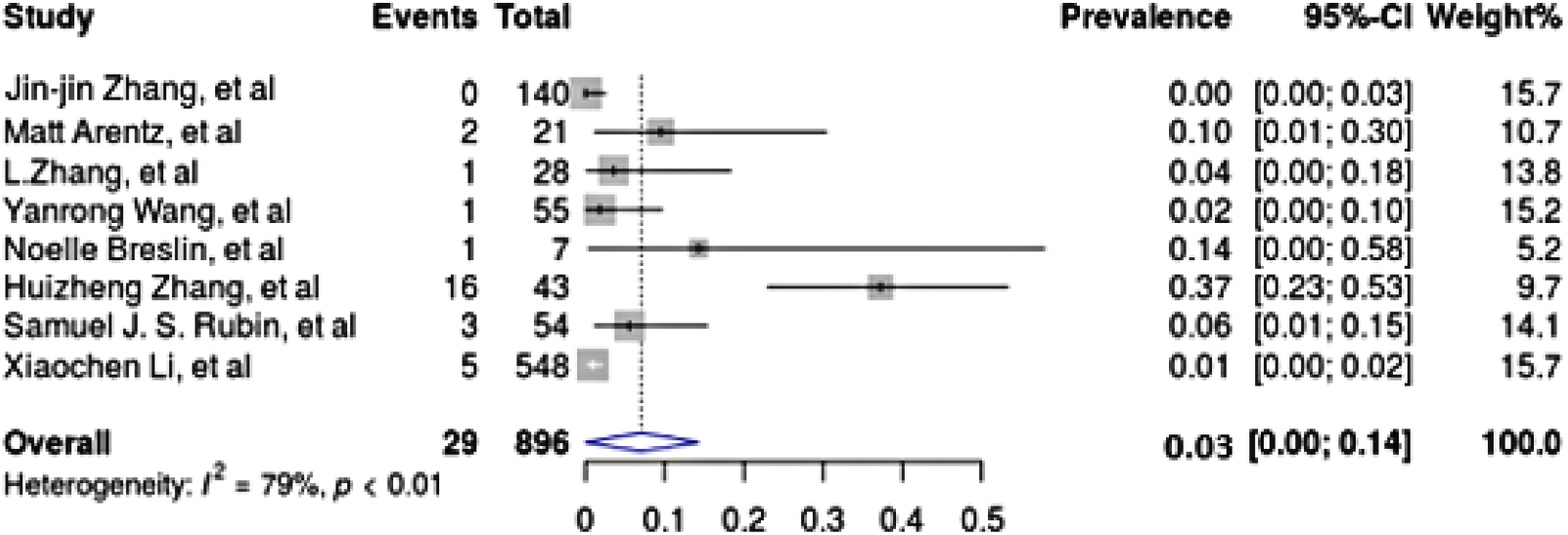
Prevalence of asthma among patients hospitalized with COVID-19.

**Fig 3:**
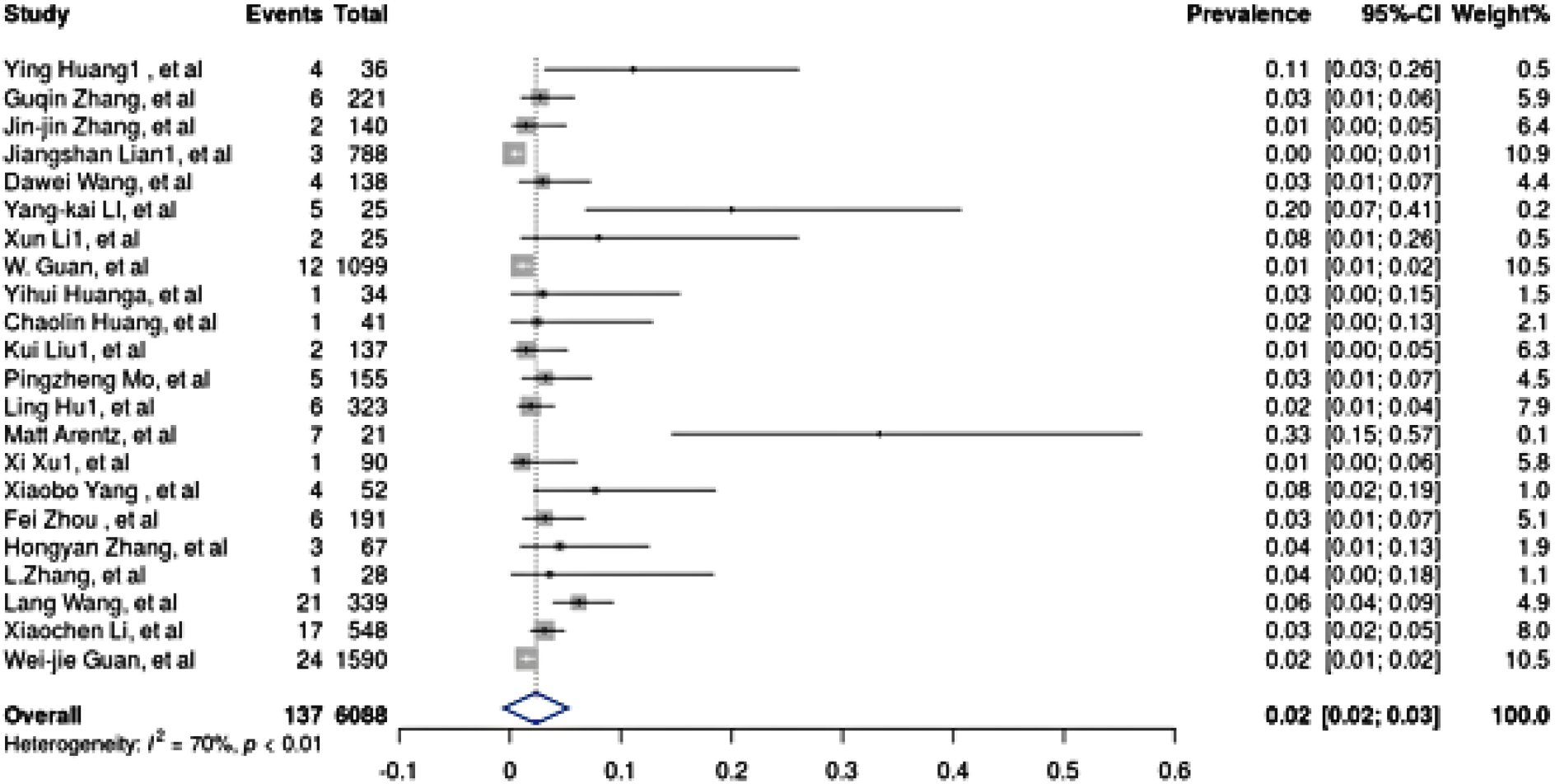
Prevalence of chronic obstructive pulmonary disease (COPD) among patients hospitalized with COVID-19.

## COPD

The pooled prevalence of COPD identified in 22 studies among patients hospitalized with COVID-19. It was estimated to be 2.2% (95% CI=0.02-0.03%).^15-17, 22-40^ Figure 3 shows the forest plot for COPD prevalence among COVID-19 patients. Our analysis identified significant presence of heterogeneity in our estimate (*I*^2^ =70%, p<0.01). The estimate of the heterogeneity variance using the statistic *I*^2^ was 70 %, value of t is 6.05 and the p-value was <0.0001, which means that a high significant publication bias was present. This is represented in of the funnel plot in Figure 3. As a result, we carry out a trim and fill method in figure 3. Here, the asymmetry outlying part of the funnel plot was trimmed off and number of studies was estimated. This estimate showed little change, which indicated influence of the publication bias was small. Egger’s test was (t = 0.66, p-value = 0.51).

## Lung Cancer

We identified 6 studies that measured prevalence of lung cancer among patients diagnosed with COVID-19.^15, 28, 38, 41-43^ In pooled analysis we found that prevalence of lung cancer among patients hospitalized with COVID-19 was 2.1% (95% CI=0.00-0.21%). Figure 4 shows the forest plot for lung cancer prevalence among COVID-19 patients. Our analysis identified significant presence of heterogeneity in our estimate (*I*^2^ = 93%, p<0.01). A significant publication bias was found in studies (t = 5.18, p = 0.01) in funnel plot see Figure 3. Consequently, we implement a trim and fill method in figure 3. These studies were used to estimate the true center of the funnel. This estimate showed little change, which indicated influence of the publication bias was small. Egger’s regression test was (t = 0.46, p = 0.66).

**Fig 4:**
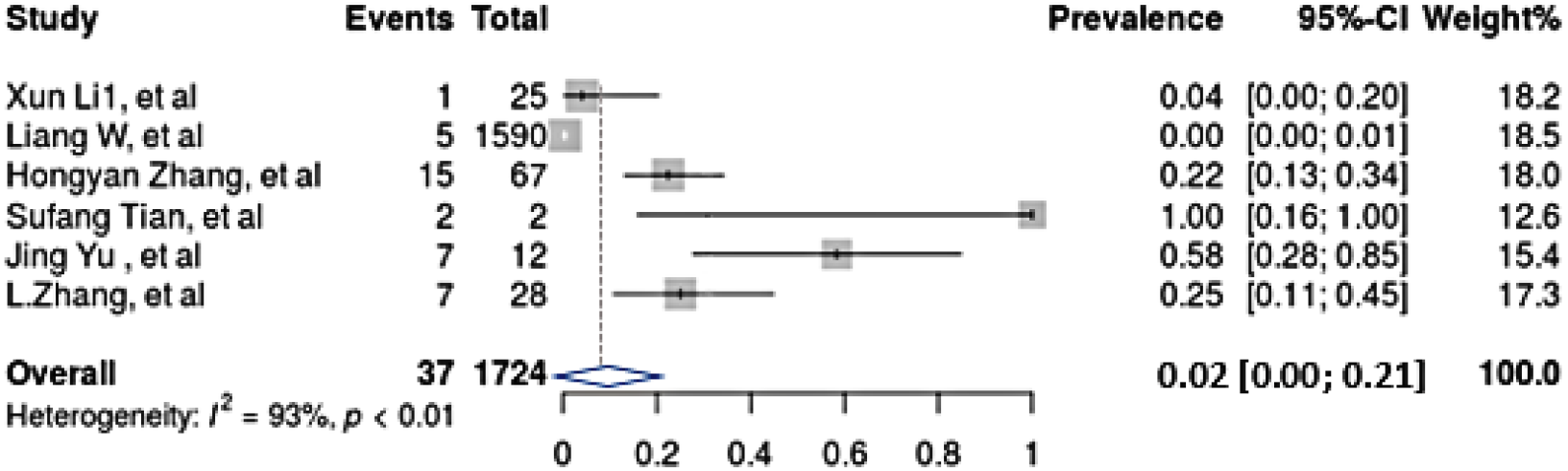
Prevalence of lung cancer among patients hospitalized with COVID-19.

## Quality Assessment

High levels of quality of the included studies in a meta-analysis are preferable. In our study, there was variable quality of the included studies. Another limitation is that the funnel plot indicated there was publication bias because there was asymmetry. Using the trim and fill method we showed that there was no change in the center of the proportion and therefore the influence of publication bias was small (Figure 5). Further, NOS assessment showed that risk of bias ranged from 4 to 7; all the included studies scored ≥ 4, which indicates low risk of bias.

**Fig 5:**
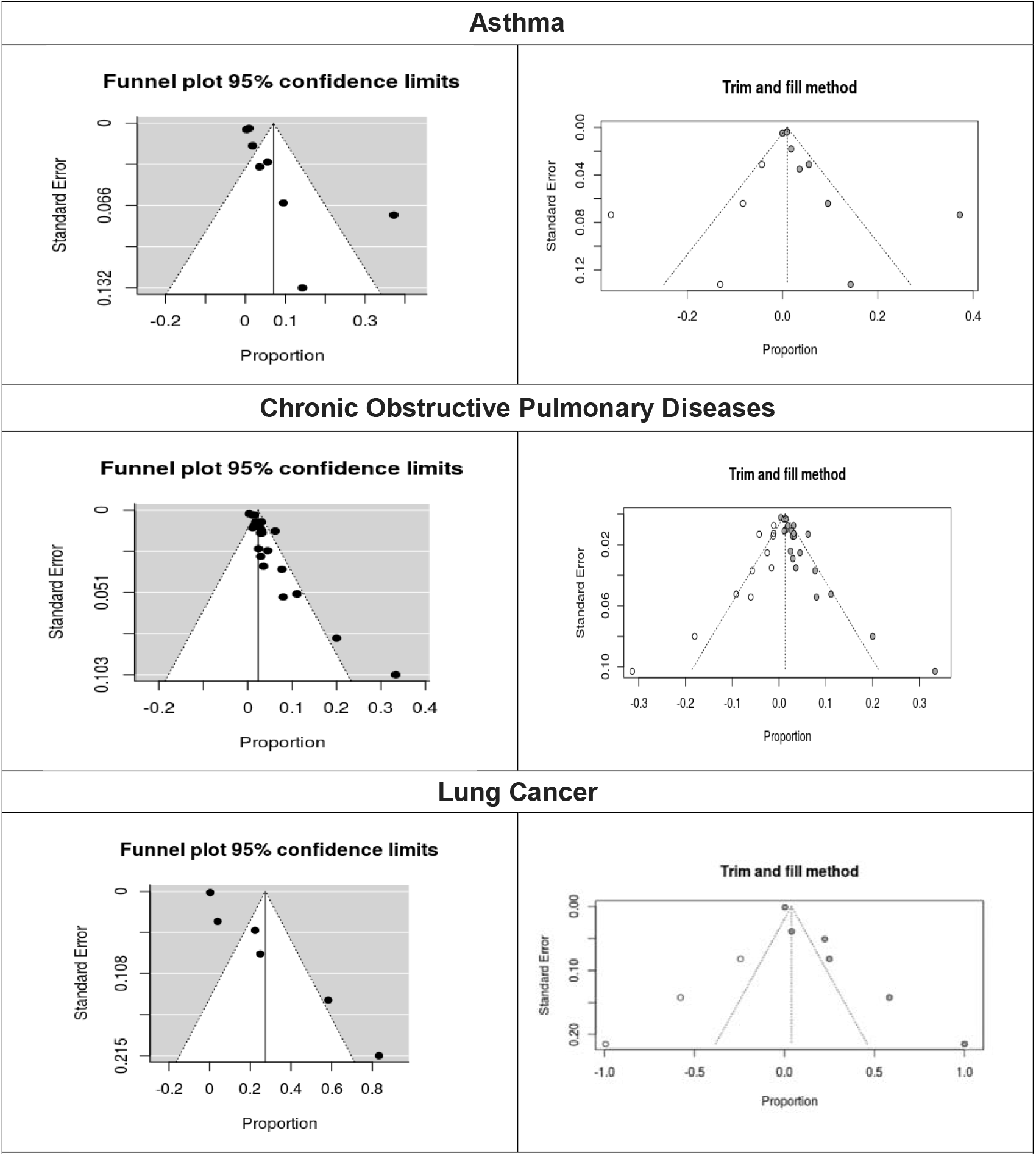
Funnel plot for meta-analysis of the prevalence of underlying diseases in COVID-19 infected cases.

## Discussion

This study helps provide and strengthen the growing evidence that comorbidities play a major role in risk profiling for at risk patients for COVID-19. Though every individual faces the risk of contracting COVID-19, many remain asymptomatic. However, the elderly and those with underlying comorbidities have shown to not only be affected by COVID-19 infection but have a more severe infection, they are more likely to face adverse outcomes, including life-threatening illnesses, use of life-saving support equipment’s including ventilators and succumb to COVID-19 related illnesses of the heart and the lung^44, 45^. Hence, individuals with underlying comorbid conditions are particularly vulnerable and should provide a high level of care including prevention and treatment to maximize benefits and minimize harms.

We found that the pooled prevalence of COPD among COVID-19 patients ranged from 2-3%. COPD remains an important sequalae of smoking associated behaviors. COPD is an obstructive pulmonary disorder that encompasses emphysema and chronic bronchitis, characterized by increasing breathlessness, bronchial inflammation, and a decline in lung function tests. Globally, COPD incidence continues to increase with smoking prevalence and age; it is now considered to be an important comorbid condition that impacts both smoking associated outcomes including lung cancer and overall quality of life.^8^ Further, prior reports on prevalence of COPD among COVID-19 patients have estimated low prevalence of COPD in range from 0.8-2%.^46, 47^ Further COPD has been shown to be associated with a nearly fourfold increase in the risk of COVID-19 infection.^44^ Also, our mortality prevalence among COPD patients was similar to rates reported previously.^46, 47^

A potential mechanism underlying higher risk of COVID-19 infection among patients with smoking-related illnesses directs towards the evidence of smoking associated upregulation of ACE-2 receptors in lower airways (type 2 pneumocytes) leading to higher rates of infection with COVID-19 among patients with underlying smoking associated comorbidities.^8^ The microbial structure of coronavirus is characterized by transmembrane (spike or S) glycoproteins namely S1 and S2.^48^ The S1 contains sites for bindings of an angiotensin-converting enzyme that provides the virus the pathway to enter into the host cells while the S2 domain is responsible for cellular infiltration.^49^ Recent studies have shown that ACE2-receptors provide a binding site for SARS-COV-2 i.e. COVID-19 with an affinity 10-20 times as compared to other coronaviruses.^35^ Further, patients with COPD also show high levels of ACE2 leading to a potentially higher risk of infection with COVID-19, mediate other potential enzymatic mechanisms, leading to lung injury, respiratory failure, and ultimately death.^50, 51^

In the pooled analysis, we found that the prevalence of asthma among patients hospitalized with COVID-19 was 3% (CI, 0-14%). Current evidence suggests that asthma remains important morbidity among patients diagnosed with COVID-19.^52^ Further in a report from New York City, the prevalence of asthma was 12.5% among patients diagnosed with COVID-19.^53^ Patients diagnosed with asthma are highly prone to infections including COVID-19 that can trigger their asthma exacerbations, thereby leading to these patients presenting to primary and emergency departments with common symptoms associated with COVID-19 including fever, dry cough and shortness of breath.^54^ Further, due to differential expression of underlying genes including ACE2 and TMPRSS2 among asthma patients based on clinical and demographic characteristics, it further warrants the need to identify the subgroup of asthma patients who might be at heightened risk of COVID-19 and develop appropriate isolation, treatment, and management strategies.^55^ Finally, patients with asthma have shown to have a weakened immune system including a deficiency and delay in lung cell interferon (IFN)-α, IFN-β and IFN-λ(7) leading to asthma exacerbations and risk of viral infections. Quantifying these cytokines might be helpful to further elucidate underlying mechanisms for progression of disease and COVID-19 and develop potential therapeutic targets.^56^ In a case report of patient who died of COVID-19 and had underlying asthma, post-mortem analysis showed mixed features of mucus plugging (hallmark of asthma) and diffuse alveolar damage (hallmark of COVID-19 pneumonia).^57^ Hence it remains imperative to manage patients with asthma with utmost care to prevent exacerbations and associated morbidity and mortality

We found that prevalence of lung cancer among patients hospitalized with COVID-19 was 2.1% (95% CI, 0.00-0.21%). Cancer predisposes an individual to a heightened risk of viral and bacterial infections due to weakened immune system. In a retrospective analysis of patients with COVID-19 and cancer in Wuhan, China, authors identified that lung cancer was the most common cancer (25%) among COVID-19 patients with cancer.^15^ Further the study found that patients with cancer were more likely to have more adverse events, including admission to ICU, use of mechanical ventilation and death.^15^ Due to the complexity of multiple factors surrounding cancer patients including patients’ clinical and demographic characteristics like age, gender, site, and stage as well as treatment and comorbidity history, certain groups recommend that baseline SARS-CoV-2 testing for all patients affected by lung cancer should be recommended and practiced.^58^ Finally, in a pathology study of 2 patients who died of lung cancer, autopsy findings identified features associated with early COVID-19 infection including edema, proteinaceous exudate, focal reactive hyperplasia of pneumocytes with patchy inflammatory cellular infiltration, and multinucleated giant cells.^42^

COVID-19 pandemic has led to an urgent need to focus on high risk patients including the elderly with underlying comorbidities. Evidence suggests that 14%–24% of patients developed pneumonitis and required hospitalization and oxygen support.^58^ Further, about 5% of patients with COVID-19 infection develop acute respiratory distress syndrome (ARDS) or sepsis-related acute organ dysfunction, requiring admission to intensive care units.^58^ Given the highly contagious nature of the virus and its affinity for lung tissue, special focus and emphasis on developing guidelines for the management of patients with underlying lung conditions remain critical to preventing disability and death.

Our study has limitations and strengths. COVID-19 is rapidly evolving globally. Hence as new literature continues to be reported, estimates of individuals with COVID-19 and chronic comorbidities might change. Further, the majority of our studies were from China, the epicenter of the COVID-19 outbreak. Hence our results might not be an adequate representation of COVID-19 cases from the global perspective. Furthermore, certain studies reported low to medium risk of bias which might be due to heterogeneity due to sample size differences, data collection procedures, differences in assessment of comorbidities or evaluation of COVID-19, and assessment of other characteristics. Our strengths include the updated use of all available literature on lung-specific comorbidities and calculating pooled estimates as compared to earlier reports.^46, 47, 59^

To conclude, our results identify key areas for future work. First, designing and implementing smoking cessation interventions on a nationwide scale to help reduce risk to high-risk smokers. Second, identifying high risk individuals in community settings with underlying lung comorbidities including those with uncontrolled COPD and asthma. Third, promoting and identifying new treatment strategies for these conditions that reduce the risk of immunosuppression, thereby reducing the risk of COVID-19 infection. Finally, isolating, tracking and tracing COVID-19 patients with lung comorbidities to prevent adverse outcomes.

In summary, our review aimed to study the prevalence and mortality associated with lung comorbidity among patients with COVID-19. It shows that the most prevalent lung comorbidity associated with COVID-19 was asthma followed by COPD and lung cancer, while the higher mortality rate was found in COPD followed by lung cancer. Future studies are needed to assess prevalence and mortality of other lung comorbidities among patients diagnosed with COVID-19.

## Data Availability

All relevant literature has been included in the manuscript.

